# Exposing racial bias in midwifery education: A content analysis of images and text in Myles Textbook for Midwives

**DOI:** 10.1101/2021.10.07.21264614

**Authors:** Mairi Harkness, Chlorice Wallace

## Abstract

**Objective:** To determine how people of different races and skin colours are represented within Myles Textbook for Midwives and whether the identified content is clinically relevant to people of all skin colours.

**Design:** Content analysis of text and images in Myles Textbook for Midwives 17th Edition, 2020

**Findings:** The images overwhelmingly depict light skinned people of White European appearance. When people of colour are shown they are more likely to be positioned in prominent imagery without specific link to the chapter topic. Descriptions of skin colour in the context of clinical assessment and/or treatment often applied mostly or solely to people with light colour skin. This included text referring to serious conditions or situations associated with severe morbidity or mortality.

**Key conclusions:** Myles Textbook for Midwives presents a light skinned White European norm and often fails to include information that is clinically relevant to the assessment and treatment of people with darker skin colours. This may lead to disparity in midwifery education and contribute to poorer outcomes for women and babies.

**Implications for practice:** Concrete efforts are required to identify and root out racial bias at all levels of midwifery education. This needs to happen alongside addressing current lack of good quality evidence required to support practice.

## Background

Race is well documented as a key factor in perinatal outcomes for women and babies. In the UK Black women are four times more likely to die during childbirth and Black babies have a 121% increased risk of stillbirth and a 50% increased risk of neonatal death when compared to babies of White ethnicity (Knight et al, 2021, Draper et al, 2018). Serious morbidity is far higher in all groups of women and babies who do not identify as White (Draper et al, 2018, Lindquist et al, 2013). The reasons for this are multiple, complex and poorly understood.

There is no doubt that racism and racial bias are present in nursing and midwifery practice and education in the UK (Lord, 2020, Horn, 2020, Burnett et al, 2020, Hunt and Richens, 1999), but it is rarely called out or openly discussed and is mostly invisible to White people, although never to Black, Asian and Minority Ethnic (BAME) colleagues (Burnett et al, 2020, p1). When racism is invisible to so many people it is often because it has become normal and ordinary and this lack of acknowledgement makes it particularly difficult to address (Stefancic and Delgado, 2010, p7, 8). Inherent racial bias has been identified within learning materials used by health care professionals including: white bodies presented as the norm in text and images; races other than White European presented as biological risk factors; and identification of concepts such as stereotyping, cosmetic bias; and fragmentation or isolation, (Sadker, 2021, Louie and Wilkes, 2018, Martin et al, 2016, Tsai et al, 2016, King and Domin, 2007, Byrne et al, 2003). The representation of white bodies as the norm and ‘othering’ of those who are not White not only reinforces power imbalance and hegemonic whiteness, it may also directly impact disparity in clinical outcomes. Louie and Wilkes (2018) note that when white bodies are normative ability to identify signs of disease in other groups may be impeded.

Stefancic and Delgado (2010, p7&8) contend that race is socially constructed and that categorising people according to race has no correspondence with biological or genetic reality. In health care settings categorising people according to race can risk misrepresenting their clinical need, with Louie and Wilkes (2018) using the example of skin cancer to highlight the importance of disentangling race and skin tone in relation to clinical risk: ‘some physicians will miss signs on dark-skinned Black patients because they do not know how to look for abnormalities. Others will miss the signs on light-skinned Black patients if they assume the ‘high risk’ skin cancer population is white’ (p41). Everett et al (2012, p7) also highlight the dangerous tendency for health care workers to ascribe characteristics based on categorisations of race rather than an individual person’s physiology and phenotype. Skin colour, as opposed to race, is an important clinical indicator and Everett et al (2012, p7) note the need for individualised care that takes this into account when assessing for jaundice, pallor, cyanosis and the blanch response, and during assessment of wounds for colour change that might indicate healing, worsening or infection. An editorial in Nurse Education Today (Burnett et al, 2020) emphasises the need for learning resources to reflect multiple ethnicities to account for differences, and similarities, in assessment and diagnosis. Menage et al (2021) describe teaching as historically skewed towards those with light skin tones leaving midwives with a knowledge gap around detection of clinical signs on darker skins. Sommer (2011) in warning against the danger of well-intentioned ‘colour blindness’ advocates for ‘colour awareness’: skin colour is relevant to health and shouldn not be ignored.

Myles Textbook for Midwives is the best-selling midwifery textbook globally. The most recent edition was published in July 2020 (Marshall and Raynor, 2020), is described as having ‘been the seminal textbook of midwifery for over 60 years.’ and as offering ‘comprehensive coverage of topics fundamental to 21^st^ century midwifery practice.’ (Elsevier, 2020). Midwifery textbooks are integral to midwifery education and practice, they not only reflect curriculum but also present implicit and explicit discourses and narratives that reveal dominant ideology and hierarchies within the profession (Harkness and Cheyne, 2019). The most recent edition of Myles Textbook for Midwives (Marshall and Raynor, 2020) was analysed in order to explore and understand its representation of race and skin colour and how that may impact clinical assessment.

## Methods

This work is a content analysis of text and images in Myles Textbook for Midwives 17^th^ Edition (Marshall and Raynor, 2020), with the aim of determining how people of different races and skin colours are represented within the textbook; and whether the identified content is clinically relevant to people of all skin colours.

### Identification & analysis of images: photographs and illustrations

A simple content analysis of the frequency and context of representation of people of different skin colours and race in images and photographs was undertaken. The analysis drew from the work of Louie and Wilkes (2018) and Martin et al (2016).

All images and photographs of people where a face is visible, and all images of an arm, head etc with visible skin were selected. Images of bone, muscle, depersonalised schematic diagrams and internal organs were excluded. The images were then categorised according to the skin colour/tone and racial group of the people represented. Race was categorised as either White or person of colour (PoC). This was determined according to observable (perceived) characteristics such as skin colour/complexion, hair texture and colour, eye colour and facial features (Roth 2012 cited by Louie and Wilkes, 2018, Martin et al, 2016). People of colour included people who are Black, Asian, Latino, Native-American and of multi-race. Skin tone was categorised as light, medium or dark using the Massey-Martin skin colour guide (cited by Louie and Wilkes, 2008) and the neonatal skin colour scale developed by Mayo-Enero et al (2020).

A simple analysis was undertaken, and descriptive statistics produced. In addition, the images were categorised according to the criteria shown in Table 1.

**Table 1:** Categorisation of images

| Category of Image | Category definition |
| --- | --- |
| Depiction | Image depicts a physical condition/phenomenon on mother or newborn. For example: cleft palate, striae gravidarum |
| Physiological | Image depicts a specific aspect of physiology. For example: fetal position in labour |
| Demonstration | Image shows demonstration of a clinical skill or activity. For example: palpation, perineal suturing |
| Illustrative | Image depicts activity directly relevant to the chapter/topic. For example: family, birth partner |
| General | Image has no specific link to the chapter or topic. For example: a mother and baby with no specific context |

### Identification & analysis of relevant text

In addition to analysis of images, a content analysis of all in-text references to skin colour made in relation to clinical assessment or treatment was undertaken. One author read the entire textbook highlighting all references to race, ethnicity and skin colour or tone. Manual reading of the textbook identified a number of words associated with skin colour and tone or used to depict a clinical description or condition strongly associated with skin colour or tone (Table 2). An electronic search was conducted to identify any additional missed text that contained reference to skin colour or tone to identify all text that describes skin colour in relation to clinical assessment or treatment.

**Table 2:**
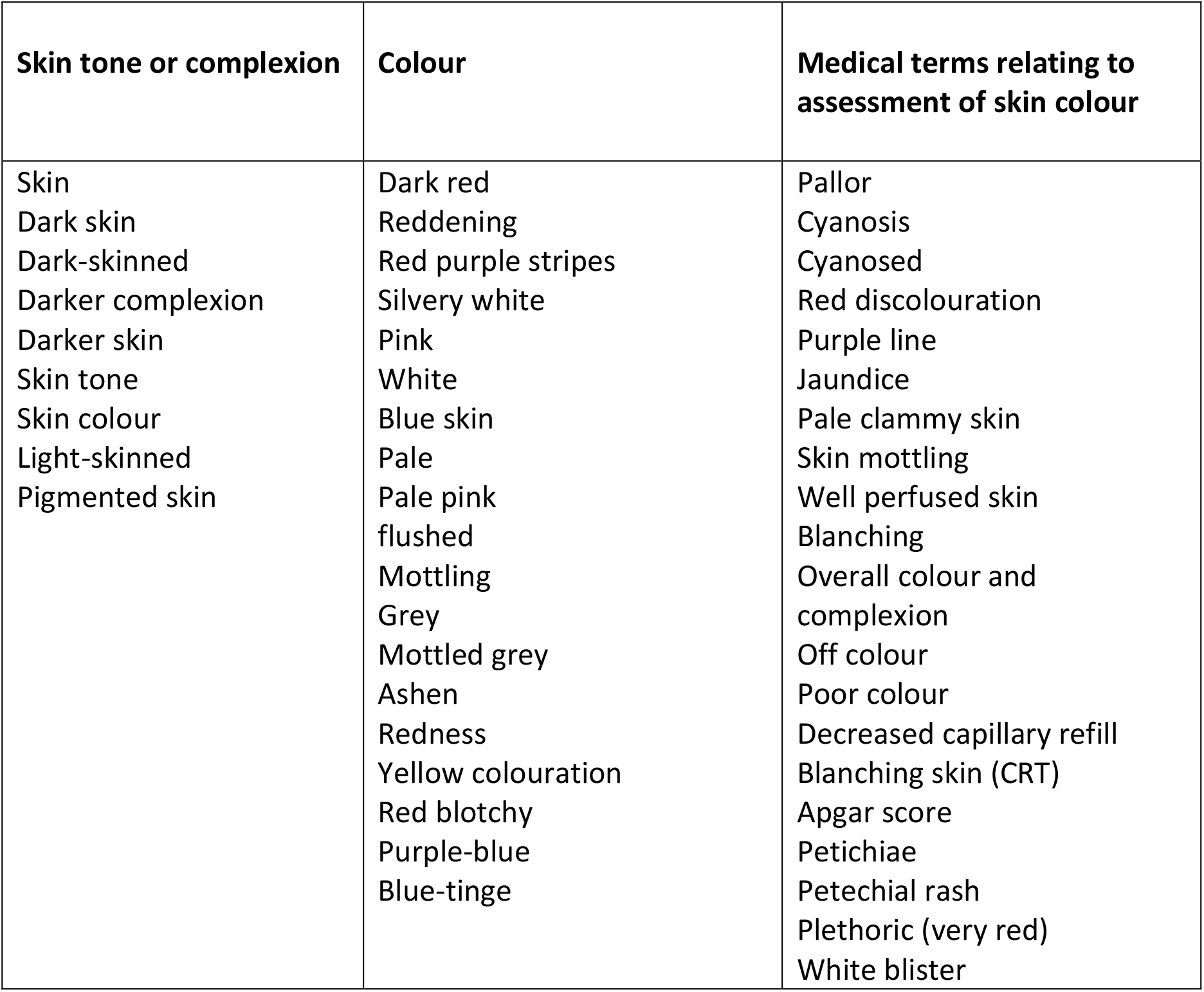
Words used to denote skin colour or clinical assessment using skin colour

| **Skin tone or complexion** | **Colour** | **Medical terms relating to assessment of skin colour** |
| --- | --- | --- |
| Skin<br>Dark skin<br>Dark-skinned<br>Darker complexion<br>Darker skin<br>Skin tone<br>Skin colour<br>Light-skinned<br>Pigmented skin | Dark red<br>Reddening<br>Red purple stripes<br>Silvery white<br>Pink<br>White<br>Blue skin<br>Pale<br>Pale pink<br>flushed<br>Mottling<br>Grey<br>Mottled grey<br>Ashen<br>Redness<br>Yellow colouration<br>Red blotchy<br>Purple-blue<br>Blue-tinge | Pallor<br>Cyanosis<br>Cyanosed<br>Red discolouration<br>Purple line<br>Jaundice<br>Pale clammy skin<br>Skin mottling<br>Well perfused skin<br>Blanching<br>Overall colour and complexion<br>Off colour<br>Poor colour<br>Decreased capillary refill<br>Blanching skin (CRT)<br>Apgar score<br>Petichiae<br>Petechial rash<br>Plethoric (very red)<br>White blister |

### Content analysis of text

All text that referred to skin colour in the context of clinical assessment and/or treatment were identified and included for analysis. The texts were then analysed from a clinical perspective to determine whether they apply to women and babies of all skin colours and what the clinical implications, if any, are if they do not.

Two midwives who work in maternity units serving diverse populations in the UK and who are familiar with clinical care of women and babies with different skin colours, categorised the text excerpts using the following guide:

*Thinking about the references to skin colour within the following extracts of text, how does this inform care of babies or mothers with different skin colours?*

1. This applies only to babies or mothers with lighter skin
2. This applies more to babies or mothers with lighter skin
3. This applies to all babies or mothers equally, regardless of their usual skin colour
4. This applies more to babies or mothers with darker skin
5. This applies only to babies or mothers with darker skin

*Does this text excerpt refer to a condition or situation that is mild, moderate or severe?*

- Mild: unlikely to cause serious morbidity
- Moderate: holds some potential for serious morbidity
- Severe: holds significant potential for serious morbidity or mortality

## Ethical Considerations

This work did not require formal ethical approval.

## Limitations

The images were categorised according to the researchers’ perceptions of the race of the people depicted and we acknowledge that that may not be the same as the race those people identify with. The concept of race is socially constructed and not based on any biological or genetic reality (Martin et al, 2016, Stefancic and Delgado, 2010) and determining someone else’s race solely from an image of them will always be subjective and imprecise. That said, the work is a necessary analysis and follows precedent of other published research.

Only one textbook was analysed and the findings do not tell us anything about other midwifery textbooks or educational materials. It is also impossible to say from this work how different people will apply their own interpretation and understanding of the text and images to clinical practice. However, Myles Textbook for Midwives, is the worlds bestselling midwifery textbook and the edition analysed was published in 2020, a time when there was heightened awareness of and focus on racial disparity in perinatal outcomes. As such, the work gives valuable insight to a prominent and important source of midwifery education that many midwives will use as a primary learning resource.

## Findings

### Content analysis of images

In total 103 illustrations depicting 262 people (adults and babies), and 67 photographs showing 108 people (adults, babies, and children) were included. The analysis found that the images used in Myles Textbook for Midwives (Marshall and Raynor, 2020) overwhelmingly represent light skinned people of White European appearance (Table 4). Among the illustrations the only people of colour depicted were all shown in just one figure, a reproduction of a World Health Organisation (WHO) poster: ‘Ten steps to successful breastfeeding’. All other illustrations depict light skinned White people. Photographs were more representative with 81% showing White people and 84% people with light coloured skin (Table 4).

**Table 4:** Representation of people of different race and skin colour in images within Myles (number of people depicted)

|  | White | Person of colour |  |
| --- | --- | --- | --- |
| Illustrations<br>(n=262) | 235 (90%) | 27 (10%)* |  |
| Photographs<br>(n=108) | 87 (81%) | 21 (19%) |  |
| All images<br>(n=370) | 322 (87%) | 48 (13%) |  |
|  | Light Skin | Medium Skin | Dark Skin |
| Illustrations<br>(n=262) | 244 (93%) | 4 (2%) | 14 (5%) |
| Photographs<br>(n=108) | 91 (84%) | 4 (4%) | 13 (12%) |
| All images<br>(n=370) | 335 (90%) | 8 (2%) | 27 (7%) |
| *All illustrations of people of colour were in one figure, reproduction of a WHO poster. |  |  |  |

It is of note that the prominent photographs used on the front and inside cover are far more diverse than those inside the textbook. One photograph on the cover was excluded as the people in the image were too small to analyse. Of the five photographs included four show people of colour, and of the 13 adults, children and babies seven (54%) are White and six (46%) are people of colour.

The analysis also classified each image according to five defined categories (Table 1), finding that the overwhelming majority of the illustrations included for analysis were used to depict clinical care and almost all were in the categories ‘physiological’ and ‘demonstration’. All images in those two categories depicted light skinned adults, babies and children with White European features (Table 5).

**Table 5:** Number and percentage of people depicted in each category of illustration

| Category of Image | White | Person of Colour |  |
| --- | --- | --- | --- |
| General | 3 (1%) | 0 |  |
| Depiction | 0 | 0 |  |
| Physiological | 135 (57%) | 0 |  |
| Demonstration | 90 (38%) | 0 |  |
| Illustrative | 7 (3%) | 27(100%)* |  |
| **Total** | 235 | 27* |  |
| Category of Image | Light Skin | Medium Skin | Dark Skin |
| General | 3 (1%) | 0 | 0 |
| Depiction | 0 | 0 | 0 |
| Physiological | 135 (55%) | 0 | 0 |
| Demonstration | 90 (37%) | 0 | 0 |
| Illustrative | 16 (7%) | 4 (100%)* | 14 (100%)* |
| **Total** | 244 | 4 * | 14 * |
| *all of these illustrations were in one figure: a reproduction of the WHO poster 'Ten steps to successful breastfeeding' |  |  |  |

Although the photographs were more diverse overall, people of colour were more likely to be depicted in an image in the ‘general’ category, that had no specific link to the chapter or topic (Table 6). Considering all photographs showing White people, 8% were in the ‘general’ category but for people of colour this was 38%.

**Table 6:** Number and percentage of people shown in each category of photograph

| Category of Image | White | Person of Colour |  |
| --- | --- | --- | --- |
| General | 7 (8%) | 8 (38%) |  |
| Depiction | 27 (31%) | 5 (24%) |  |
| Physiological | 0 | 0 |  |
| Demonstration | 44( 51%) | 8 (38%) |  |
| Illustrative | 9 (10%) | 0 |  |
| **Total** | 87 | 21 |  |
| Category of Image | Light Skin | Medium Skin | Dark Skin |
| General | 11 (12%) | 0 | 4 (31%) |
| Depiction | 27 (30%) | 2 (50%) | 3 (23%) |
| Physiological | 0 | 0 | 0 |
| Demonstration | 44 (48%) | 2 (50%) | 6 (46%) |
| Illustrative | 9 (10%) | 0 | 0 |
| **Total** | 91 | 4 | 13 |

### Content analysis of in-text references to skin colour

All text that referred to skin colour in the context of describing clinical assessment and/or treatment were identified and analysis undertaken to determine whether they were clinically relevant to people of all skin colours. In total 62 pieces of text were included for analysis.

The analysis found that the majority of references to text excerpts were either in category 1: ‘this applies only to babies or mothers with light skin’, or category 2: ‘this applies more to babies or mothers with light skin’: 48/62 (84%) for reviewer one, and 35/62 (57%) for reviewer two. One text excerpt was categorised by one of the reviewers as being category 4: ‘this applies more to babies or mothers with darker skin’and the remaining text excerpts were assessed as being category 3: ‘this applies to all babies and mothers regardless of their skin colour/tone’ (Table 7).

**Table 7:**
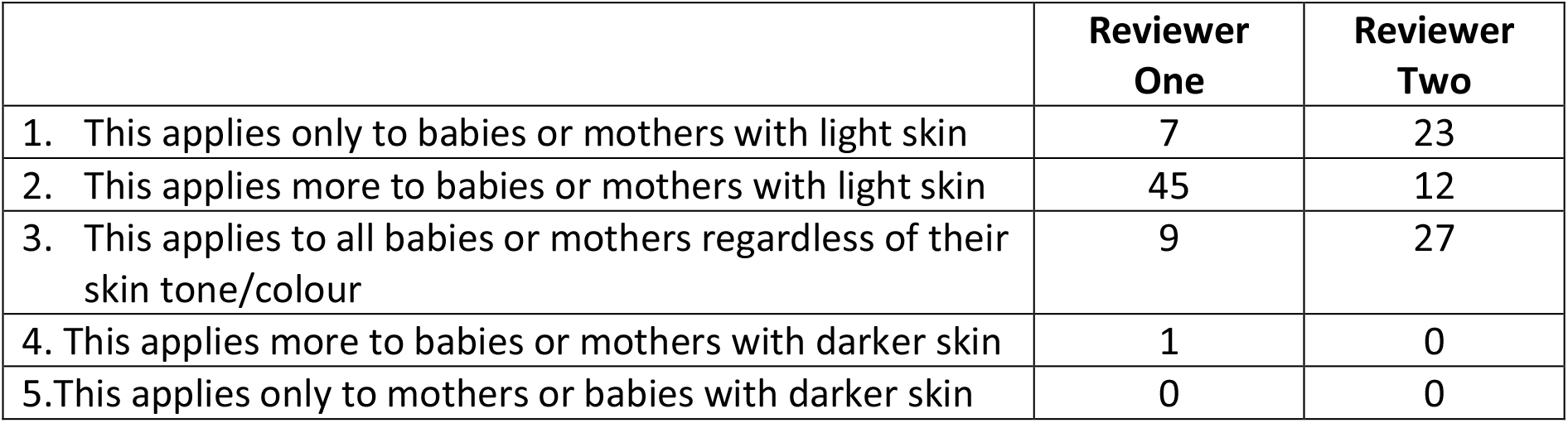
Categorisation of the text excerpts by reviewers one and two.

Most of the text excerpts were categorised as referring to conditions that were moderate: ‘holds some potential for serious morbidity’, or severe: ‘holds significant potential for serious morbidity or mortality’ (73% for reviewer one and 60% for reviewer two). Of the 41 text excerpts where the reviewers agreed that the condition or situation could be described as ‘moderate’ or ‘severe’, 32 (78%) were also categorised as applying only or more to babies or mothers with light skin (categories 1 and 2).

## Discussion

This work found that the images within Myles Textbook for Midwives (Marshall and Raynor, 2020) overwhelmingly depict people who are of light skinned White European appearance, and that when people of colour are represented they are more likely to feature prominently but less likely to be included in images that depict a clinical skill or situation directly relevant to the topic under discussion.

The ethnicity of the UK population varies greatly across its countries and regions: in Scotland 96% of people identify as White, in London 44.9% do (Scotland’s Census, 2021, UK Government, 2018). Myles Textbook for Midwives is published in the UK and sold internationally yet the images in the textbook do not represent the diversity in race and skin colour of people who will use the book or the people that they are learning about.

Sadker (2021) describes ‘cosmetic bias’ as a form of bias where there is different representation in prominent imagery than the overall textbook. This gives the illusion of equity but suggests that minimal efforts have been made to address diversity throughout. The positioning of people of colour in prominent images, particularly on the front and inside cover, indicates superficial attempt to address the books general failure to provide representative imagery in the illustrations and photography.

Analysis of the text mirrored analysis of the images in finding that information about clinical assessment focuses on people with light coloured skin. The combined findings demonstrate uneven representation with light skinned White people presented as the ‘typical’ norm on which teaching is based. This not only negates the experience of people outside the dominant White category, evidence also suggests that lack of diversity and uneven representation in educational resources may impact patient care delivery and contribute to racial inequality in healthcare experience, treatment and outcomes (Louie and Wilkes, 2018, Martin et al, 2016, Byrne, 2003). Lack of diversity within textbooks can reinforce assumptions about the ‘typical patient’ and Louie and Wilkes (2018, p41) note that when white bodies are normative ability to identify signs of disease in other racial groups may be impeded, resulting in diagnostic inequities.

The majority of text excerpts were categorised by both reviewers as applying mostly or only to people with lighter skin. However, the two reviewers did not always agree in their assessment of individual pieces of text. That two experienced clinical midwives differed in their understanding of text in relation to assessment of skin colour is an important finding in itself and led to further examination, discussion and consideration of the texts in the context of available evidence.

Much of the information provided by the textbook in relation to skin colour is highly subjective. Skin colour and tone varies across a large spectrum. Signs of jaundice, cyanosis and pallor rely on an assessment of skin colour but are recognised as having different characteristic appearance in different ethnic groups and there is conflicting evidence around how best to recognise them (Stephen et al, 2021, Kanji et al, 2017, Szabo et al, 2004).

Midwives must understand differences in usual colour of healthy skin and difference in signs that indicate deviation from normal in order to provide safe and appropriate care to all, yet this is rarely discussed explicitly within the text. Many of the text excerpts directed the reader to assess ‘colour’ without providing any other information about what that colour should be. For example:

> ‘*and the woman’s overall colour and complexion* {referring to signs of wellbeing}’.

Ex 14, p721)

> ‘if the baby has poor colour and muscle tone {resuscitation of health baby at birth}’

Ex 67, p853

Byrne (2003) describes failure of nursing learning resources to represent diverse groups by using a framework of Minnich’s ‘errors basic to dominant tradition’. These errors include ‘faulty generalisation’ when one specific group is represented but the content is generalised to all people, and ‘circular reasoning’ when a norm or ideal is based on an exclusive category, usually defined by a dominant (p277). Using phrases such as ‘assess colour’ or ‘poor colour’ become problematic when the reader’s main point of reference throughout the textbook is a White person with light coloured skin.

Other excerpts were more explicitly problematic in their failure to describe conditions in a way that would allow the reader to identify clinical problems in people with darker skin colour.

> ‘Sign: Appearance (colour): Score: 0-pale or blue; 1 -body pink; extremities blue; 2-completely pink’ {Apgar score}

Ex 64, p849

> ‘Skin: Gelatinous, red, translucent; Smooth, pink, visible veins; Cracking, pink areas, rare veins’ {assessment of healthy low birth weight baby -signs of physical maturity: skin}

Ex 71, p865

Visual assessment of skin colour is subjective, with this subjectivity further complicated by variation of usual skin colour across racial/ethnic groups. The particle size, shape and location of melanin is the most significant factor in relation to overall colour, and the more melanin that is clustered nearer the surface of the skin the darker skin will appear (Everett et al, 2012). The extent to which skin appears ‘pink’, ‘red’, ‘blue’, ‘yellow’, ‘grey’ or ‘white’ varies dependent on a person’s usual skin colour and the subjective assessment of the person examining them.

Many commonly used medical words and the concepts behind them are founded on assessment of light coloured skin: cyanosis from Greek ‘Kyaneos’ meaning blue, and jaundice from the French ‘jaune’ meaning yellow. The pieces of text that discuss assessment and treatment based on skin colour or tone, often in relation to life threatening situations such as resuscitation or major haemorrhage, rarely provided information about what ‘colour’ or ‘cyanosis’ or ‘pallor’ or ‘jaundice’ look like if a person does not have light coloured skin.

When usual skin colour was mentioned references were brief, of limited value and positioned people with darker skin as different. They often reverted to describing a light skinned norm within the section or chapter. For example:

> ‘Clinical recognition and assessment of jaundice can be difficult, especially in babies with dark skin tones. In the UK, the use of a transcutaneous bilirubinometer (TCB) is recommended to measure the bilirubin level’

Ex 51, p932

> ‘Unconjugated bilirubin is fat soluble and will deposit in subcutaneous fat, which makes the skin appear yellow’

Ex 53, p933

The evidence base around the use of TCB for people with darker skin is contradictory. Although their use is considered more effective than visual clinical assessment, not all types of TCB are effective for all skin colours (Szabo et al, 2004). Historically people from BAME backgrounds have been under represented in clinical and health research, limiting the validity and generalisability of studies that ostensibly apply to the whole population (Redwood and Gill, 2013) This contributes to lack of understanding around the need for different approaches to clinical assessment and a deficit of evidence available to inform the care of people with darker skin.

If understanding around difference in skin colour is not included in midwifery educational resources midwives become reliant on individual experience to develop their understanding. Disparity in effective learning opportunities, given the large difference in ethnic mix across the countries and regions of the UK, puts at risk the provision of safe, effective and equitable care for all women and babies.

Reluctance to identify and discuss difference in skin colour may arise from well-intentioned colour blindness, in the mistaken belief that equity of care means ignoring skin colour. However, equitable care requires the opposite: colour awareness, explicit acknowledgment and explanation of difference (Sommers, 2011). Structural racism describes institutional practices that benefit White people and disadvantage people of colour, it is present throughout society and midwifery education is no exception. When education focuses on a light skinned norm clinical disadvantage for those with darker skin manifests in many ways; from recognition of perineal trauma, signs of domestic abuse and wound healing, through to identifying jaundice, cyanosis or pallor (Everett et al, 2012, Sommers, 2011). It is this structural racism, not race, that puts people at risk (Crear-Perry, 2021, Hardeman et al, 2016).

## Conclusion

Black people and people of colour are at greatly increased risk of harmful perinatal outcomes. The reasons for this are multiple and complex and must be identified and addressed. This work found that Myles Textbook for Midwives, the most popular and widely used midwifery textbook in the world, presents white bodies as the norm in text and images and fails to provide information that is relevant to clinical assessment of mothers and babies with darker skin.

Structural racism is ubiquitous throughout society and its manifestation in midwifery education may be a contributing factor to the current disparity in outcomes. Local efforts to decolonialise curricula and learning materials are to be welcomed, however much more is required. Concrete efforts to identify and root out racial bias at all levels of midwifery education is necessary, and this must happen alongside addressing the current lack of good quality evidence to support practice.

Using the terms racism and racial bias may feel uncomfortable, particularly when individuals are in fact committed to treating people equally, but if a problem is not identified and named it cannot be addressed. Positive change is only possible when underpinned by understanding that it is racism, not race, that puts women and babies at risk.

## Data Availability

All data produced in the present work are contained in the manuscript

## Data Availability

All data produced in the present work are contained in the manuscript

## Data Availability

All data produced in the present work are contained in the manuscript

## Acknowledgements

The authors would like to thank Nahida Hanif and Memuna Sowe for their substantial contribution to this work. We would also like to thank Professor Helen Cheyne for her support and advice.

## Notes

### Competing Interest Statement

The authors have declared no competing interest.

### Funding Statement

No external funding was received for this work

